# Increased Tryptophan, But Not Increased Glucose Metabolism, Predict Resistance of Pembrolizumab in Stage III/IV Melanoma

**DOI:** 10.1101/2022.12.06.22283165

**Authors:** Jorge D. Oldan, Benjamin C. Giglio, Eric Smith, Weiling Zhao, Deeanna M. Bouchard, Marija Ivanovic, Yueh Z. Lee, Frances A. Collichio, Michael Meyers, Diana E. Wallack, Amber Abernethy-Leinwand, Patricia K. Long, Dimitri G. Trembath, Paul B. Googe, Madeline H. Kowalski, Anastasia Ivanova, Jennifer Ezzell, Nana Feinberg, Nancy E. Thomas, Terence Z. Wong, David W. Ollila, Zibo Li, Stergios J. Moschos

**Author notes:** **Corresponding Author:** Dr. Stergios J. Moschos. Equally contributed to the work presented herein (Zibo Li, Stergios J. Moschos).

## Abstract

Clinical trials of combined IDO/PD1 blockade in metastatic melanoma (MM) failed to show additional clinical benefit compared to PD1-alone inhibition. We reasoned that a tryptophan-metabolizing pathway other than the kynurenine one is essential. We immunohistochemically stained tissues along the nevus-to-MM progression pathway for tryptophan-metabolizing enzymes (TMEs; TPH1, TPH2, TDO2, IDO1) and the tryptophan transporter, LAT1. We assessed tryptophan and glucose metabolism by performing baseline C11-labeled α-methyl tryptophan (C11-AMT) and fluorodeoxyglucose (FDG) PET imaging of tumor lesions in a prospective clinical trial of pembrolizumab in MM (clinicaltrials.gov, NCT03089606). We found higher protein expression of all TMEs and LAT1 in melanoma cells than tumor-infiltrating lymphocytes (TILs) within MM tumors (n=68). Melanoma cell-specific TPH1 and LAT1 expression are significantly anti-correlated with TIL presence in MM. High melanoma cell-specific LAT1 and low IDO1 expression were associated with worse overall survival (OS) in MM. Exploratory optimal cut-point survival analysis of pretreatment ‘high’ vs. ‘low’ C11-AMT SUV_max_ of the hottest tumor lesion per patient revealed that the ‘low’ C11-AMT SUV_max_ was associated with longer progression-free and OS in our clinical trial (n=26). We saw no such trends with pretreatment FDG PET SUV_max_. Treatment of melanoma cell lines with telotristat, a TPH1 inhibitor, increased IDO expression and kynurenine production in addition to suppression of serotonin production. High melanoma tryptophan metabolism is a poor predictor of pembrolizumab response and an adverse prognostic factor. Serotoninergic but not kynurenine pathway activation may be significant. Melanoma cells outcompete adjacent TILs, eventually depriving the latter of an essential amino acid.

## Introduction

L-tryptophan (Trp) is an essential amino acid in translation and protein synthesis. However, the majority (>95%) of Trp serves as a biochemical precursor of metabolites with critical physiologic roles (reviewed in ^1^) in gut homeostasis, immunity, and neuronal function. Under normal physiologic conditions, 95% of the absorbed Trp is metabolized via the kynurenine pathway by the action of indolamine 2,3-dioxygenases 1 (IDO1/IDO2) and Trp 2,3-dioxygenase (TDO2). The resultant kynurenines are biologically active metabolites involved in inflammation, immunoregulation, excitatory neurotransmission, and gut homeostasis. Dysregulation of kynurenine metabolism can lead to various conditions, such as cancer, diabetes, mood, and psychiatric disorders (reviewed in ^2^).

Increased IDO1 and TDO2 expression have been previously reported across various malignancies. In addition, increased kynurenines have broad immunoregulatory properties and can function as oncometabolites.^3^ Despite promising results in early clinical studies of combined IDO1 and PD1 inhibition in metastatic melanoma (MM),^4^ randomized clinical studies failed to show more significant clinical benefit than PD1 inhibition alone.^5^ Trp metabolism in melanoma may be more complex than initially thought.

Apart from the kynurenine pathway, the role of the serotoninergic pathway, the alternative pathway to Trp metabolism, is less understood. Less than 5% of the absorbed Trp is metabolized via the Trp hydroxylase 1 (TPH1) and TPH2 towards serotonin (5HT)/melatonin. Serotonin plays an important physiologic role as a neurotransmitter, growth factor, angiokine, and inflammatory modulator. In cancer, serotonin has growth-stimulatory, migration/invasion, progression, and angiogenic properties (reviewed in ^6^). Interestingly, high expression of TPH1 across different immune cell subsets can induce an immune-tolerant state.^7^ Finally, melanoma cells can express TPHs and produce serotonin/melatonin.^8^

The lack of clinical benefit from combined IDO1 and PD1 inhibition in MM, the importance of Trp in immune system functioning, and the potential for non-kynurenine pathway-specific mechanisms of resistance of PD1 inhibitors led us to investigate the protein expression of all four Trp metabolizing enzymes (TMEs) and their transporter, LAT1, in melanoma. Furthermore, we have assessed Trp metabolism in patients with PD1 inhibitor-naïve stage IIIB-IV melanoma in a prospective clinical study of pembrolizumab (LCCC1531; clinicaltrials.gov, NCT03089606). We hypothesized that metabolic dysregulation within the tumor depletes Trp via a pathway less dependent on IDO1 expression and activity. The resultant immune dysfunction via intratumoral Trp depletion may contribute to a lesser clinical benefit from PD1 inhibitors and possibly decreased overall survival (OS).

## Patients and Methods

### The University of North Carolina at Chapel Hill (UNC-CH) Melanocyte/Melanoma cell line Array (CLA), the Biomax Normal Skin/Benign Nevus/Primary Melanoma Tissue Microarray (TMA), and the UNC-CH 09-1737 Metastatic Melanoma TMA

We have used: (a) our previously described CLA that consists of normal human melanocytes (NHM) and 38 melanoma cell lines for immunocytochemistry (ICC).^9^ (b) commercially available TMAs that contain normal skin, benign nevi, and primary melanomas (SK181 and ME1002b; US Biomax, Inc., Derwood, MD) for immunohistochemistry (IHC). (c) our previously described TMA that consists of MM tissues collected from patients who underwent standard-of-care (SOC) surgery for stage III/IV cutaneous melanoma at UNC-CH.^9^

### The LCCC1531 Trial

#### Patients

We enrolled adult patients with biopsiable stage IIIB-IV melanoma who had not previously received PD1 inhibitors. The study was approved by UNC-CH’s Institutional Review Board (IRB). Written informed consent was obtained from all patients in compliance with the IRB’s regulations. **Supplementary Material** (*<u>Patients and Methods</u>*) describes major eligibility criteria.

**Supplementary Material** *(<u>Supplementary Figure 1</u>)* outlines the sequence and timing of key tests and procedures. Patients underwent SOC positron emission tomography (PET) scan using fluorodeoxyglucose (FDG) co-registered with computerized tomography (CT) scan of the neck, chest, abdomen, and pelvis using intravenous (IV) contrast. After investigators identified a measurable (and biopsiable) tumor lesion(s) by Response Evaluation Criteria in Solid Tumors (RECIST) v1.1 criteria, patients underwent an α-[^11^C]-methyl-L-tryptophan (C11-AMT) PET scan of the tumor lesion(s) of interest at the UNC-CH’s Biomedical Research Imaging Center. A research tumor biopsy was performed after completing the C11-AMT PET scan and before pembrolizumab initiation. Enrolled subjects were treated with pembrolizumab, 200 mg flat dose IV, administered over 30 min every three weeks until progression, intolerable toxicity, or completion of up to four pembrolizumab infusions.

**Supplementary Material** (<u>Patients and Methods</u>) describes details regarding the C11-AMT PET scan protocol acquisition, image analysis, and patient monitoring during and after study completion.

#### Mandatory Baseline (pretreatment) Tumor Tissue Biopsies

Before pembrolizumab treatment, we collected mandatory research biopsies from tumor tissues previously imaged by both FDG-PET and C11-AMT PET. Representative 5μm-thick tissue sections were stained with hematoxylin and eosin (H&E) to assess for tumor content, necrosis, and density of TILs by expert pathologists (DGT, PBG).

### Tissue Procurement, Single-Color Immunohistochemistry, and Digital/Manual Pathology Scoring

We immunohistochemically stained representative 5μm-thick sections from previously described TMAs, CLAs, and the LCCC1531 trial’s research biopsies with commercially available antibodies against TPH1, TPH2, TDO2, IDO1, and LAT1. Normal human tissues and commercially available normal TMAs were antibody-stained according to the organ-specific pattern for each protein (BN501a, BN126, BN117a, US Biomax, Inc. Rockville MD). In addition, we stained LCCC1531 tissue sections for the two principal hexokinase enzyme isoforms involved in the first glycolysis step, hexokinase (HK) 1 and HK2, and the glucose transporter 1 (GLUT1, SLC2A1). **Supplementary Material** (*<u>Supplementary Table 2</u>*) describes antibodies, dilutions, and conditions for each single-color IHC assay.

The CLA and TMA slides were digitally imaged at 20x magnification using the Aperio ScanScope XT (Leica Biosystems). Images were analyzed using Aperio’s Color Deconvolution Algorithm (Leica Biosystems). We presented the results as a histologic score (HScore, 0-300).^10^

Analysis of the normal skin/benign nevi/primary melanoma TMA slides (SK181, ME1002b), the metastatic melanoma 09-1737 TMAs, and the LCCC1531 tissue slides were manually performed by expert pathologists. Briefly, we analyzed the status of TILs (present, absent) and expression/abundance/cell distribution of the Trp and glucose metabolism-related proteins. For scoring, we used the 0, 1+ (<25% of cells with a membrane/cytoplasmic stain), 2+ (25-80% of cells with a membrane/cytoplasmic stain), 3+ (>80% of cells with membrane/cytoplasmic stain) semiquantitative scale to describe the staining intensity and percentage of positive melanoma cells.^10^ If tissue cores also included TILs, we yielded a separate 0 to 3+ score for staining intensity and percentage of positive TILs.

### In Vitro Studies

Details on statistical analysis are shown in **Supplementary Material**.

### Statistical Analysis

Details on statistical analysis are shown in **Supplementary Material**.

## Results

### Expression of TME in NHMs, Melanoma Cell Lines, Normal Skin, Nevi, and Primary Cutaneous Melanomas

We assessed protein expression of the TMEs and LAT1 in NHMs (n=3), melanoma cell lines (n=41), normal human skin, nevi (n=7), and primary melanomas (n=51) by single-color ICC/IHC (**Figure 1A). Supplementary Material** (*<u>Supplementary Table 3</u>*) shows the protein expression of TMEs in NHM and melanoma cell lines. We saw no significant differences in protein expression in any of the five proteins among NHMs and all other melanoma cell lines (**Figure 1B**). However, among the melanoma cell lines, TPH1 and LAT1 expression were significantly higher in *BRAFV600*-mutant (n=22) compared to *RAS*-mutant (n=11) melanoma cell lines (unpaired *t*-test with Bonferroni adjusted *p*-value <0.01 for both**; Figure 1B**).

**Figure 1.**
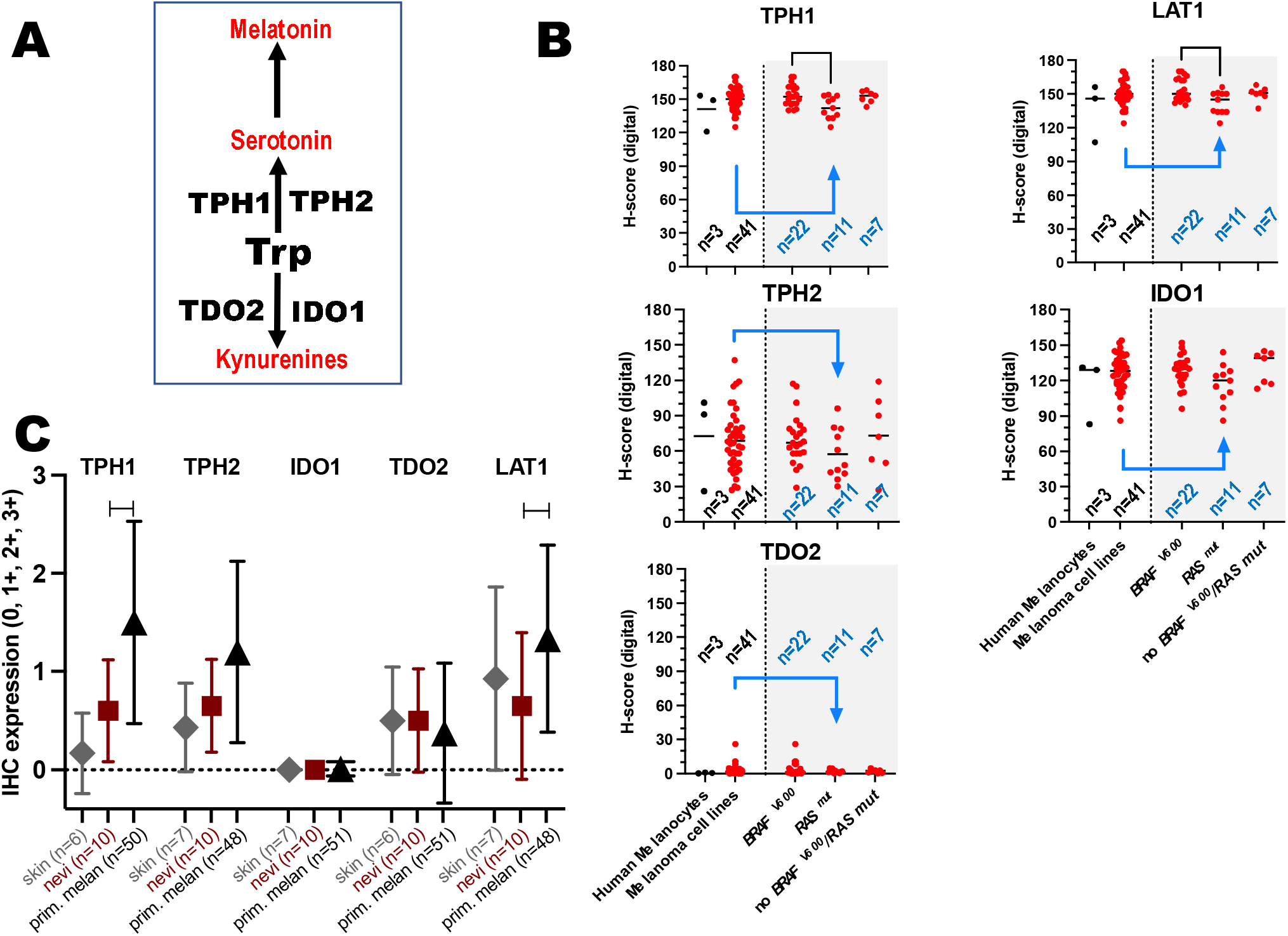
Protein expression of the four TMEs and LAT1 in melanoma. **A**. The enzymatic role of each TME is shown in the first rate-limiting enzymatic step of Trp metabolism. **B**. Expression (digital H-score) in NHMs (black, n=3) and melanoma cell lines (red, n=41). Within melanoma cell lines, expression is further shown (blue arrow, light grey region) according to the mutation status for *BRAFV600* and *RAS* mutations. **C**. Expression (semiquantitative 0-3 score) in normal skin (n=7), nevi (n=10), and primary melanomas (n=51). See, **Supplementary File** (<u>Patients and Methods</u>) for histopathologic details in the analysis. Asterisks indicate significant differences in protein expression between nevi and primary melanomas. **, p<0.01; * p <0.05.

**Figure 1C** shows that the expression of TPH1, TPH2, and TDO2 was low-to-absent in normal epithelial keratinocytes and nevi. IDO1 expression was absent in normal epithelial keratinocytes, whereas LAT1 expression was low in both. Again, compared to nevi, TPH1 and LAT1 expression in primary melanomas was significantly higher in primary melanoma and did not significantly change for TPH2 and TDO2. IDO1 expression remained absent in primary melanomas.

### Expression of the TMEs in Stage III/IV Melanoma (09-1737 cohort)

We assessed protein expression of the TMEs and LAT1 in eighty-eight tumor tissues from 87 patients by single-color IHC. Patient characteristics are shown in **Supplementary Material** (*<u>Supplementary Table 4</u>*). In tumor cores with present TILs, expression of all five proteins was significantly higher in melanoma cells than in TILs (paired *t*-test *p*-value for all five proteins was <10^−8^). Among the expression of the four TMEs in melanoma cells, irrespective of TIL status, IDO1 was the least expressed enzyme. In contrast, expression of TPH1 and TPH2 was the highest (paired *t*-test for the comparison between each of TPH1, TPH2, and TDO2 enzymes with IDO1 with Bonferroni adjustment of *p*-value was <0.001) (**Figure 2)**. Our results suggest that in contrast to nevi or earlier melanoma stages, protein expression of TMEs from both serotonergic and kynurenine pathways is upregulated in MM; in fact, the melanoma cell-specific expression of TPH1 and TPH2 was higher than the corresponding expression of IDO1 and TDO2. However, within tumors, melanoma cells express the four TMEs and LAT1 significantly higher than TILs.

**Figure 2.**
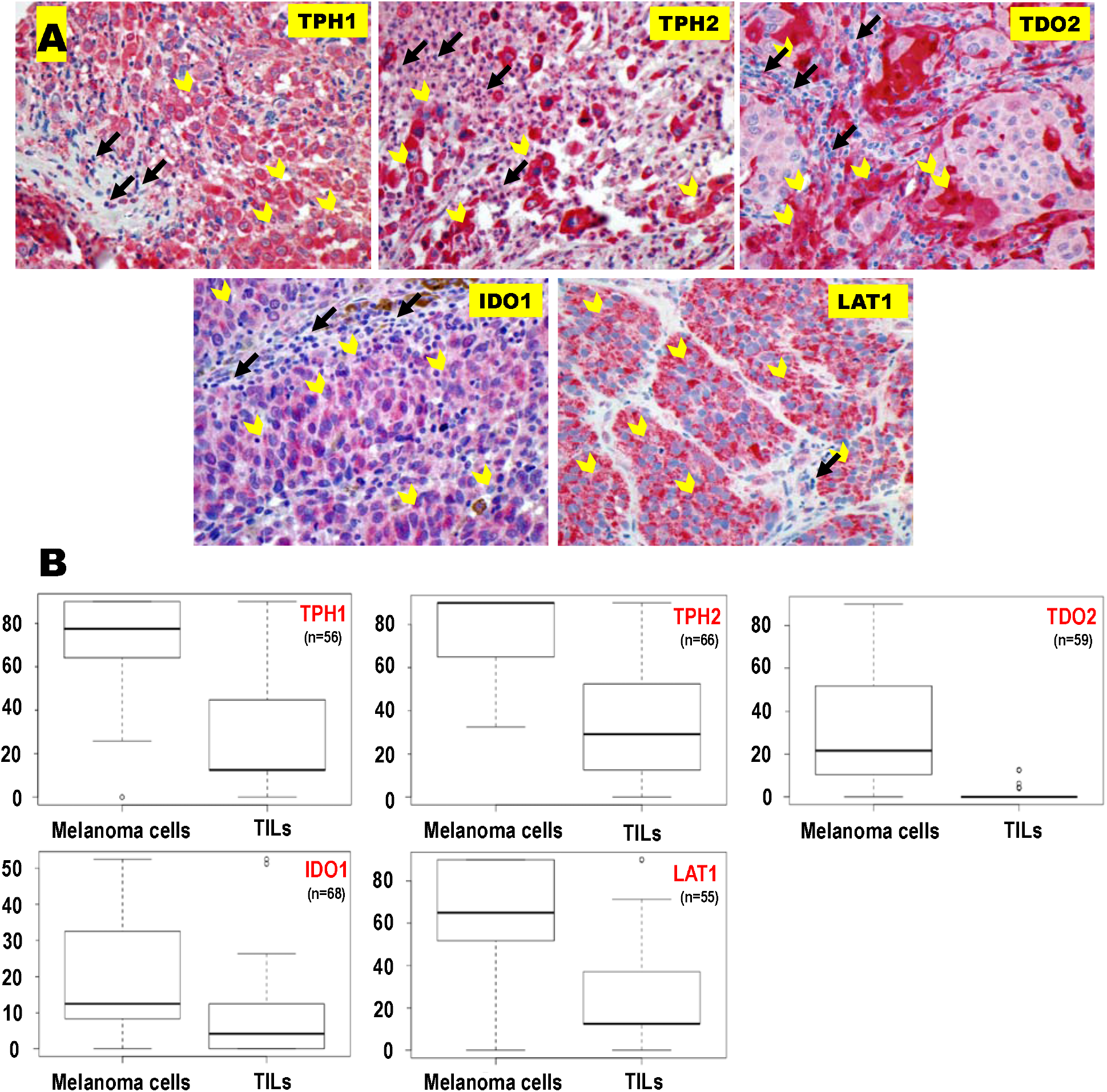
Expression of the TMEs and LAT1 in melanoma samples from stage III-IV melanoma patients. **A**. Representative images (20x magnification) were obtained from representative tumors having TILs to show differences in protein expression between melanoma cells (open yellow arrows) and TILs (black arrows). **B**. Boxplots show average protein expression of the proteins scored separately for melanoma cells and TILs. Only samples that had at least one value for expression of each protein in both melanoma cells and TILs for a given sample are shown.

We then hypothesized that if the expression of the TMEs and LAT1 is higher in melanoma cells than TILs, the former may overwhelmingly take up and clear Trp from the tumor microenvironment. Again, we found that increased expression of TPH1 and LAT1 in melanoma cells was significantly associated with reduced TIL presence (**Supplementary Material**, [*<u>Supplementary Table 5</u>]*). The estimated odds ratio (OR) of TIL presence comparing a sample with a one-unit higher TPH1 expression to a sample with lower TPH1 expression was 0.982 (95% confidence interval (95CI) [0.967, 0.998], *p*=0.030). Furthermore, the estimated OR of TIL presence for a one-unit increase in LAT1 expression was 0.984 (95CI [0.970, 0.998], *p*=0.023). The association tests between TIL presence and expression for the other three enzymes were inconclusive.

### Prognostic Significance of Melanoma Cell-specific Protein Expression of TMEs in Stage III/IV Melanoma (09-1737 cohort)

Given that TPH1 and LAT1 expression in melanoma cells negatively correlates with TIL presence, we questioned whether the melanoma cell-specific expression of TMEs and LAT1 correlates with melanoma-specific OS in stage III/IV melanoma. The median follow-up of 09-1737 patients between stage III/IV specimen collection and the last follow-up was 15 months (range 1-168 months). A Cox proportional hazard model that included the average protein expression of TMEs and LAT1 and was adjusted for age, sex, and stage showed that the estimated hazard ratios (HR) for a unit increase in LAT1 and IDO1 were 1.018 (95CI [1.004-1.032], *p*=0.016), and 0.979 (95CI [0.960-0.999], *p*=0.028), respectively (**Supplementary Material**, [*<u>Supplementary Table 6]</u>*). Our results suggest that increased melanoma cell-specific expression of LAT1 is associated with a worse OS, whereas melanoma cell-specific IDO1 expression was associated with a more prolonged OS. The tests of association of TPH1, TPH2, and TDO2 expression with OS were inconclusive.

### Theragnostic Significance of Trp and Glucose In Vivo `Metabolism in Metastatic PD1 Inhibitor-Naïve Melanoma (LCCC1531 study)

A higher protein expression of certain TMEs and LAT1 does not *per se* reflect higher metabolism and/or Trp uptake by the tumor. To assess Trp metabolism and uptake by patient tumors in real-time, we performed PET imaging using C11-AMT. By methylating Trp in its α-position, AMT can still be taken up by cells via the LAT1 transporter and metabolized by all four TMEs; however, AMT can no longer participate in downstream intracellular protein synthesis steps (**Supplementary Material**, *<u>Supplementary Figure 2</u>*). We were also interested in comparing the theragnostic utility of C11-AMT PET with the ‘gold standard’ FDG PET.

We enrolled 26 subjects between June 2017 and November 2020 at UNC-CH who underwent C11-AMT PET, FDG PET, baseline research tumor biopsies, and received at least one pembrolizumab infusion as part of the study. **Table 1** shows baseline patient characteristics. The majority were males (77%) who had cutaneous (69%) and AJCC stage IV melanoma (69%). Only four patients had prior non-surgical treatment for melanoma; three had previously received adjuvant ipilimumab, and one had received talimogene laherperepvec (T-VEC).

**Table 1.** Patient characteristics from the LCCC1531 clinical trial (n=26).

|  |  |
| --- | --- |
| **Age** (yrs; median, range) | 63 (30,90) |
| 18-64 | 14 |
| ≥65 | 12 |
| **Sex (%)** |  |
| Male | 20 (77) |
| Female | 6 (33) |
| **Melanoma Subtypes (%)** |  |
| Cutaneous | 18 (69) |
| Acral | 2 (8) |
| Mucosal | 2 (8) |
| Ocular | 1 (4) |
| Unknown Primary | 3 (11) |
| ***BRAFV600E* mutation** |  |
| Yes | 3 |
| No | 15 |
| Unknown | 8 |
| **AJCC staging (v7) at Original Diagnosis** |  |
| IIIB | 1 (4) |
| IIIC | 6 (23) |
| IIID | 1 (4) |
| M1a | 3 (11) |
| M1b | 7 (27) |
| M1c | 8 (31) |
| **Prior Cancer Therapies (number of patients, %)** |  |
| No | 23 (88) |
| Yes (adjuvant ipilimumab only) | 3 (12) |
| **ECOG Performance Status at Screening (%)** |  |
| 0 | 19 (73) |
| 1 | 7 (27) |

The median duration of pembrolizumab treatment was 5.2 months (range 0.6-31.3+ months). **Supplementary Material** (*<u>Supplementary Table 7)</u>* shows AEs that occurred in more than one study subject during the study. **Figure 3** shows the swimmer’s plots of all 26 patients in the study. Twenty of the 26 patients continued their care at UNC-CH after completing the 12-week study protocol; the remaining six received pembrolizumab treatment and follow-up radiographic imaging by community medical oncologists. All patients had radiographic imaging every three months for the first two years, every six months for the third and fourth year, and annually after that. Seven (27%) and five (19%) patients developed complete and partial radiographic responses, respectively, whereas two patients (8%) had stable disease as the best antitumor response (patients 16 and 24). Two of the partial responders elected to undergo radical lymph node dissection (patients 7 and 25); they were therefore censored for the PFS endpoint. Twelve patients (46%) progressed while on pembrolizumab; seven progressed before receiving four pembrolizumab infusions. The median PFS was 7.4 months (range 0.6-52.6+ months).

**Figure 3.**
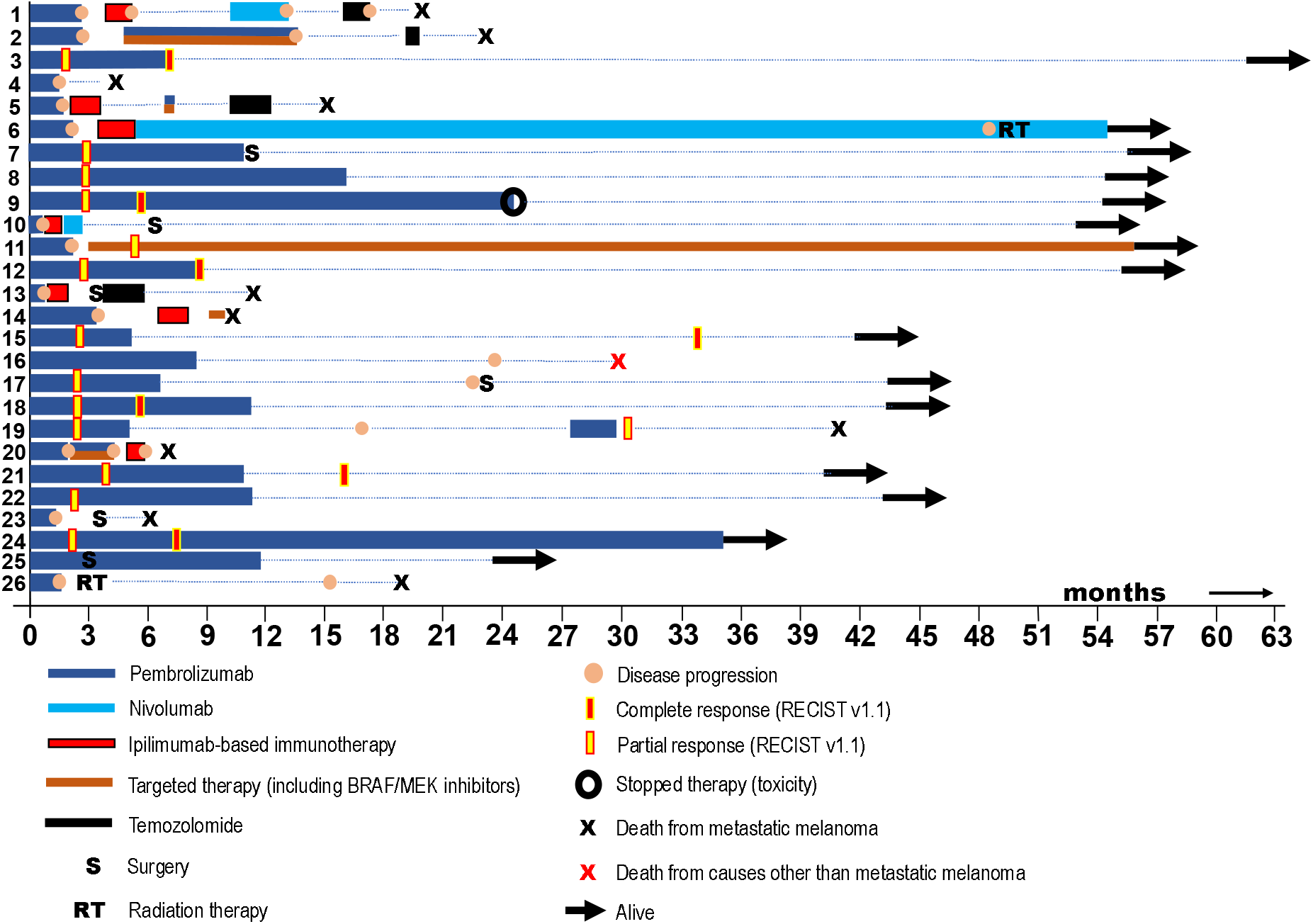
Swimmer’s plots of patients from the LCCC1531 trial.

As of November 2022, ten patients had died from MM. One additional patient with MM died from complications from COVID-19 infection (patient 16); he was therefore censored for the melanoma-specific OS endpoint. Six patients are alive with MM. The median OS was 40.8 months (range 4.3-61.7+ months).

### Theragnostic Significance of Baseline Trp and Glucose Metabolism in Metastatic PD1 Inhibitor-Naïve Melanoma (LCCC1531 study)

Sixty-one tumor lesions from all 26 patients were imaged by C11-AMT PET and FDG-PET imaging and a CT scan with IV contrast before pembrolizumab treatment. The majority of imaged tumor lesions at baseline were melanoma-infiltrated lymph nodes (n=33, 52%), followed by subcutaneous nodules (n=22, 35%), followed by liver metastases (n=7, 11%). **Figure 4, panel A**, shows baseline C11-AMT PET SUV_max_ values across all 61 tumor lesions for all 26 patients. The median C11-AMT SUV_max_ was 6.3 (range 1.1,23). **Figure 4, panel B**, shows baseline FDG PET SUV_max_ values across all 61 tumor lesions from all patients, according to tumor volume and FDG PET SUV_max_ changes following 12 weeks of pembrolizumab or earlier if progression. The median FDG SUV_max_ was 8.5 (range 2.1-22). There was a significant and moderate correlation between baseline tumor volume and SUV_max_ for both FDG and C11-AMT PET scans (Spearman rank ρ=0.53, p<0.0001 and ρ=0.45, p=0.0003, respectively). In addition, there was a significant and moderate correlation between baseline C11-AMT SUV_max_ and baseline FDG PET SUV_max_ (Spearman ρ=0.53, p<0.0001, **Figure 4, panel C**). **Figure 4, panel D**, shows representative images of individual tumor lesions using both PET tracers 52 out of 61 tumor lesions were also imaged with FDG PET after 12 weeks of pembrolizumab or earlier if progression. SUV_max_ increased following pembrolizumab treatment in 18 of the 20 growing tumor lesions (Wilcoxon matched-pairs signed rank test p=0.0005) and decreased following pembrolizumab treatment in 30 out of 31 shrinking tumor lesions (Wilcoxon p<0.0001). In contrast, the SUV_max_ of the stable lesion increased following pembrolizumab treatment.

**Figure 4.**
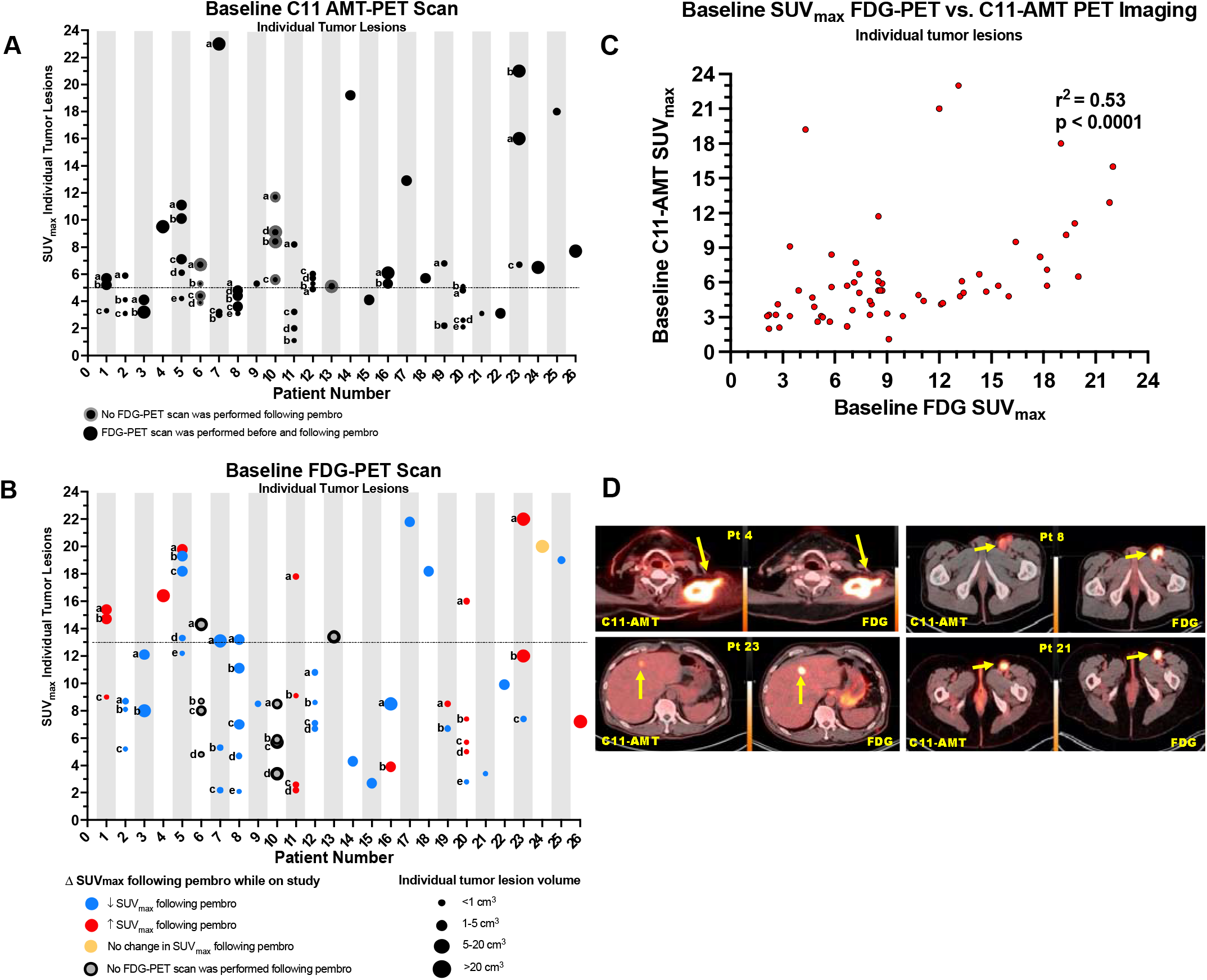
SUV_max_ of tumor lesions imaged by C11-AMT (panel A) and FDG PET scan (panel B) prior to pembrolizumab treatment (LCCC1531 trial). SUV_max_ values are shown the ratio of the voxel with the highest radiotracer concentration compared to a “background region” in close proximity to the tumor. Horizontal bars indicate conventional cutoff values for ‘high’ vs. low ‘low’ baseline SUV_max_ (optimal cutpoint analysis, see text). **C**. Correlation between baseline SUV_max_ in C11-AMT and FDG PET of tumor lesions. **D**. Representative images of tumor lesions from patients (pt). Each panel shows sections of the same tumor imaged with C11-AMT (left) and FDG PET scan (right). Patients 4 and 23 progressed whereas patients 8 and 21 responded to treatment.

We performed Cox analysis to assess the value of baseline C11-AMT PET and FDG PET in predicting clinical benefit from pembrolizumab and OS. For the PFS endpoint, the estimated coefficient for the baseline SUV_max_ of the hottest lesion per patient by C11-AMT PET imaging was 1.06 (p=0.15), whereas the corresponding coefficient for FDG-PET imaging was 1.05 (p=0.31). We then performed exploratory *post-hoc* optimal cutpoint analysis of ‘high’ vs. ‘low’ SUV_max_ values for baseline C11-AMT PET and FDG PET imaging. By setting SUV_max_ cutoff value of 5 for C11-AMT PET and 13 for FDG PET for ‘high’ vs. ‘low’ SUV_max_, we found that patients with ‘low’ SUV_max_ C11-AMT PET trended to have either stable disease, partial response, or complete response by RECIST v1.1 criteria (Fisher’s exact test p=0.08). However, patients with ‘low’ baseline SUV_max_ of the hottest tumor lesion per patient by C11-AMT PET scan had a significantly prolonged PFS and OS benefit compared to patients with ‘high’ baseline C11-AMT PET scan. The baseline SUV_max_ FDG PET scan was not associated with either PFS or OS (**Figure 5**).

**Figure 5.**
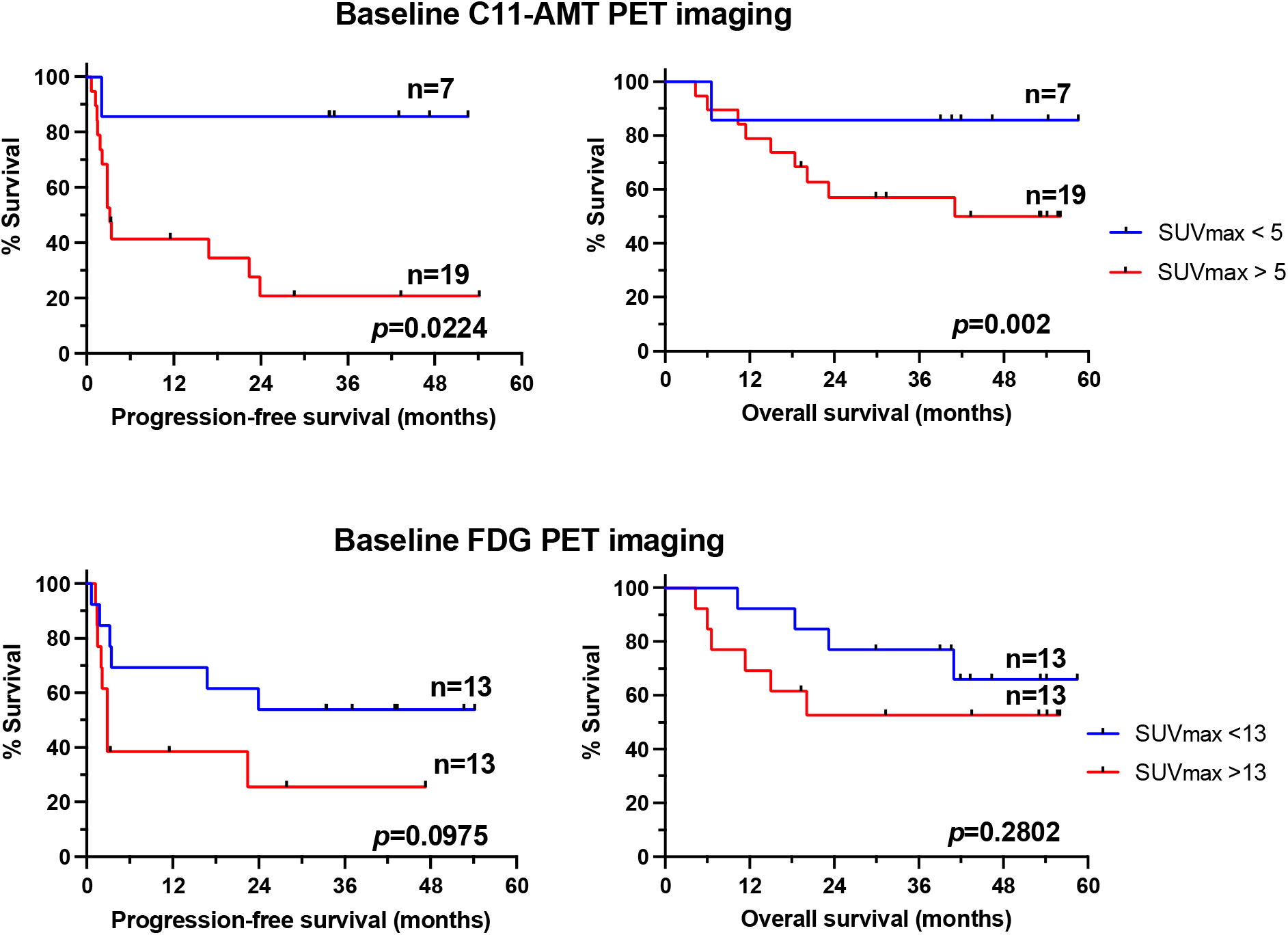
Theragnostic significance of baseline SUV_max_ using C11-AMT PET and FDG PET imaging in metastatic PD1 inhibitor refractory melanoma using optimal cutpoint *post-hoc* analysis. The hottest tumor lesion from each patient was only included in the analysis.

### Correlative Analysis of Baseline Tumor Tissue in Metastatic PD1 inhibitor-naïve Melanoma (LCCC1531 study)

Eighteen of the 26 mandatory research baseline tumor biopsies had a sufficient amount of viable melanoma. Fortunately, six of the eight patients whose research biopsies did not have viable melanoma had archived metastatic tumor specimens collected prior to study enrollment from previous SOC surgical procedures. Thus, 18 patients who had tumors imaged by both FDG-PET and C11-AMT PET also had subsequent research tumor biopsies with viable melanoma; these tumors were available for the correlation analysis between melanoma-specific IDO expression and baseline SUV_max_ parameters. Twenty-four pretreatment tumor specimens (archived plus research) were available for other tumor tissue correlative analysis.

We found that high expression of IDO1 by melanoma cells trended to significantly correlate with low C11-AMT SUV_max_ values of these tumors (Kendall tau rank correlation -0.34, p=0.08). We found no significant correlation between IDO1 expression and FDG PET SUV_max_ values (−0.20, p=0.30). In addition, there were no significant differences in either PFS or OS between patients bearing tumors with high (2+, 3+) vs. absent/low (0, 1+) TILs by H&E analysis (data not shown). We then compared the protein expression for each protein involved in the Trp and glucose metabolism between melanoma cells and TILs, if present. After adjusting the critical *p*-value for multiple comparisons using the Bonferroni correction (p=0.0064), the expression of TPH1, TDO2, LAT1, and HK2 by melanoma cells was significantly higher than the corresponding expression by TILs (**Supplementary Material**, *<u>Supplementary Figure 3</u>*). The expression (high vs. low/absent) of none of the proteins involved in Trp and glucose metabolism was associated with PFS, OS, baseline FDG PET SUV_max_, and baseline C11-AMT PET SUV_max_ of the hottest tumor lesion per patient. Finally, we explored the correlation among the proteins involved in Trp, glucose metabolism, and TILs. Among the highest significant correlations were seen between TPH1 and LAT1 expression by melanoma and TILs (Spearman rank correlation coefficient ρ=+0.81 and +0.83, respectively; *p*<0.0001 and 0.003, respectively), between HK1 expression in TILs and TIL density (ρ=0.81, *p*=0.004) and between TDO2 expression in melanoma cells and TIL density (ρ=0.62, *p*=0.003).

### Treatment of Melanoma Cell Lines with Telotristat

The high expression of TPH1 in primary and MM, its higher expression by melanoma cells compared to neighboring TILs, and the reduced abundance of TILs in melanomas bearing a high expression of TPH1 prompted us to investigate the effect of pharmacologic inhibition of TPH1 in melanoma cell lines *in vitro*. Treatment of SK-MEL-2 and MeWo cell lines with increasing concentrations of telotristat etiprate, a TPH1 inhibitor, for 72 hours did not significantly affect cell growth or cell death. However, telotristat treatment significantly increased IDO protein expression in melanoma cells, whereas TPH1 expression remained relatively unaltered. Furthermore, the effects of telotristat on TPH1 and IDO1 expression in melanoma cells were associated with significant changes in serotonin (reduced) and kynurenine (increased) levels cell line culture medium (**Figure 6**). These results may highlight metabolic flexibility in melanoma cells, such that pharmacologic inhibition of the serotonin pathway may lead to a compensatory increase in the kynurenine pathway in melanoma cells.

**Figure 6.**
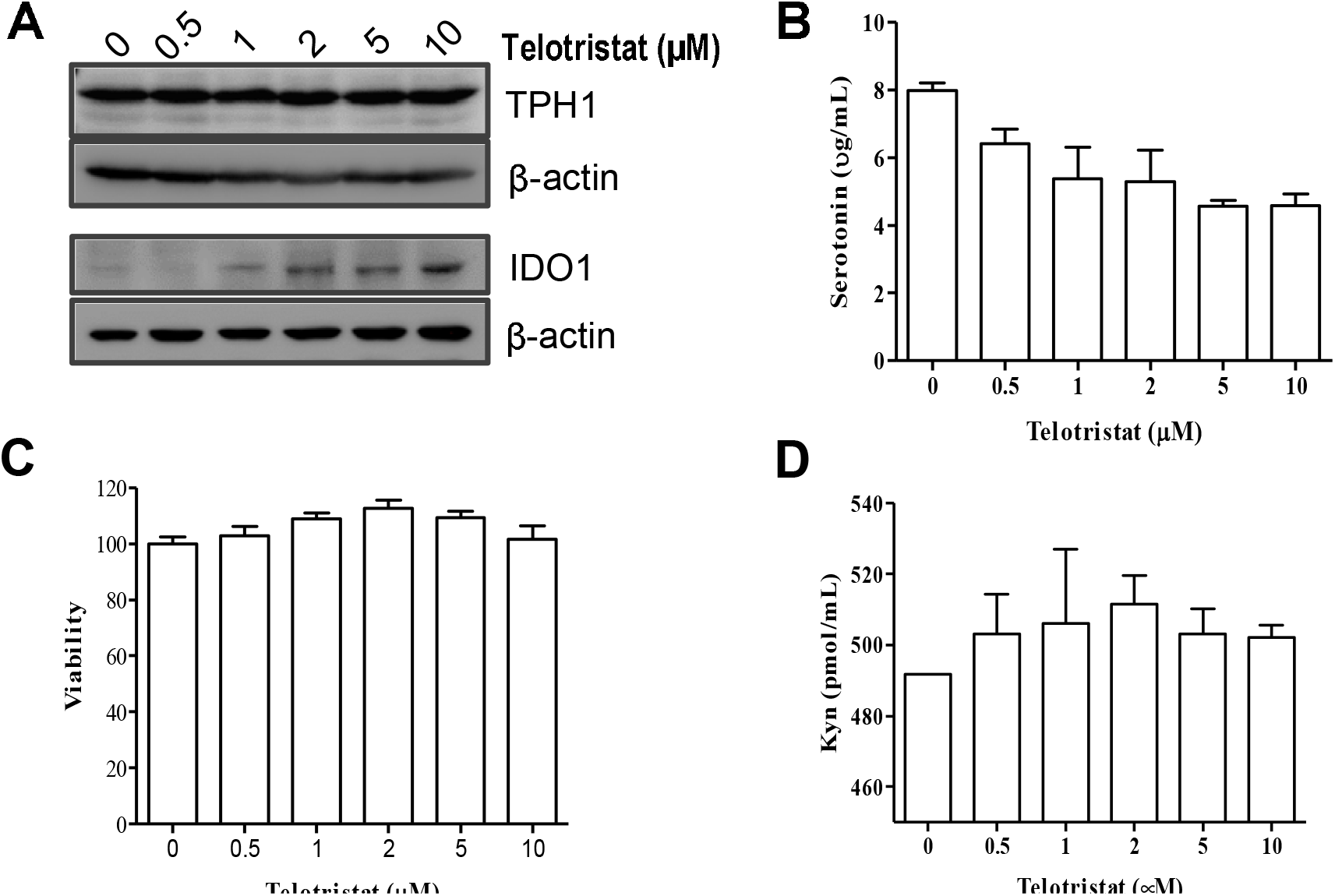
Treatment of SK-MEL-2 melanoma cells with telotristat. **A**. Immunoblot analysis of cell lysates 24 hours post treatment initiation. Experiment was performed in triplicates. **B**. Cell viability using a MTT assay 72 hours following treatment initiation. Experiment was performed in duplicates. **C & D**. ELISPOT assay for 5HT/serotonin and Kyn 24 hours following post treatment initiation. Experiment was performed in duplicates.

## Discussion

In this study, we show that MM exhibits dysregulation in Trp metabolism, similar to other solid tumors.^11,12^ Nevertheless, this is the first prospective clinical study to report that high Trp imaging is associated with shorter clinical benefit from pembrolizumab in PD1 inhibitor-naïve MM and shorter OS in a *post-hoc* optimal cut-point exploratory survival analysis. In fact, baseline C11-AMT PET imaging exhibited a better predictive value for clinical benefit to pembrolizumab than FDG PET imaging in MM.

How can the metabolism of an amino acid such as Trp have a better predictive role in clinical benefit from pembrolizumab but not glucose? Trp is an essential amino acid for mammalian cells and vital for anabolic processes in melanoma and immune cells.^1^ On the other hand, a high density of tumor-infiltrating CD8+ cells at baseline has been associated with clinical response to PD1 inhibitors in melanoma.^13^ Our IHC analysis across MM samples shows that the expression of all TMEs and LAT1 is significantly higher in melanoma cells than in TILs. We hypothesize that within this competitive tumor microenvironment, melanoma cells have a greater capacity for uptake and metabolism of Trp than adjacent TILs. Therefore, Trp depletion via metabolic competition between the melanoma cells and TILs eventually deprives lymphocytes of an amino acid necessary for proliferation.^14^ Disabled immune cells are thus forced to switch from an activated/anabolic state to a dormant/catabolic state and struggle to survive.^15^

We noticed a significant positive correlation between SUV_max_ in baseline C11-AMT and FDG PET imaging for individual tumor lesions from the LCCC1531 study. Given the significant correlation of each SUV_max_ with individual tumor volumes, we must assume that in vivo metabolism for both Trp and glucose is principally driven by melanoma cells themselves. Nevertheless, only approximately 50% of the SUV_max_ variability value in C11-AMT could be predicted by the SUV_max_ in FDG PET, suggesting that our findings cannot be solely explained by a non-specific rev up of global tumor metabolism in melanoma cells. In line with this, we were surprised to find low-to-moderate expression levels of GLUT1 in both melanoma cells and TILs, contrasting the consistently high levels of LAT1 in both cell populations. The redundancy of glucose transport may alternatively explain this discrepancy via other glucose transporters that we did not measure in this study. Alternatively, we postulate that while Trp is an essential nutrient for any cell, normal or malignant, glucose is not because glucose can be produced via different intracellular metabolic pathways, especially by the metabolically more flexible melanoma cells. Besides, we have previously shown that glycolysis is not melanoma cells’ principle metabolic pathway.^16^

Given the lack of higher clinical benefit from combined IDO1 and PD1 inhibition in MM,^5^ we sought to better understand the relative contribution of the kynurenine and serotonin/melatonin pathway in the overall Trp metabolism within tumors, given that C11-AMT is not specific for one pathway over the other. We, therefore, performed an extensive IHC analysis of cell lines and tissue specimens across all stages of the nevus-melanoma progression pathway for TMEs and LAT1. To our surprise, we identified increased TPH1 and LAT1 along the nevus-melanoma progression pathway, whereas the expression of TDO2 and IDO1 was relatively unchanged. Although upregulation of an enzyme(s) does not automatically correspond to a higher enzymatic activity, our finding that TPH1 and LAT1 were significantly higher in *BRAFV600*-mutant as opposed to *RAS*-mutant melanoma cell lines suggests that oncogenic mutations such as *BRAFV600E*, may, among other mechanisms, contribute to the metabolic reprogramming of melanoma cells by selectively upregulating serotonin/melatonin but not kynurenines. In addition, in our clinical study C11-AMT SUV_max_ of the biopsied lesion trended to anti-correlate with melanoma-specific IDO1 expression, again suggesting that the high C11-AMT SUV_max_ signal could possibly be driven by non-IDO1-associated Trp metabolizing pathways. In support of the importance of the serotoninergic pathway, a recent metabolomic analysis of sera collected from patients with IIIC/IV MM and sex-/age-matched controls showed not only significant alterations in Trp metabolism in MM but also higher serum serotonin, but not kynurenine, levels.^17^ Finally, a single remote case report showed increased intratumoral levels of serotonin in melanoma tumors.^18^ These results suggest that upregulation of the serotonin/melatonin but not the kynurenine pathway may account for the higher Trp metabolism in melanoma cells.

We have herein shown that low baseline C11-AMT PET scan SUV_max_ of the hottest tumor from the LCCC1531 patients is associated with a prolonged OS in a post hoc optimal cutpoint exploratory survival analysis; this is in line with our findings from a multivariate analysis of prognostic factors from the 09-1707 patient cohort, which showed that high LAT1 expression by melanoma cells is an adverse prognostic factor. Interestingly, high expression of IDO1 by melanoma cells was a favorable prognostic factor in the 09-1707 patient cohort, perhaps because IDO1 is upregulated in response to a pro-inflammatory environment (IFNγ, TNFα). The latter signifies the poorly characterized (pro-tumorigenic vs. antitumor) role of serotonin/melatonin in cancer.^6^

Telotristat is the first FDA-approved TPH1 inhibitor for patients with carcinoid syndrome.^19^ Interestingly, in syngeneic colorectal and pancreatic tumors, telotristat augmented effects of PD-1 checkpoint blockade and increased tumor infiltrating CD8+ cells by suppressing serotonin-mediated upregulation of PD-L1 in tumor cells.^20^ We found that telotristat suppressed serotonin production by melanoma cells without significant effects in melanoma cell proliferation. Interestingly, however, telotristat increased expression of IDO1 in melanoma cell lines, which may account for the increased kynurenin concentrations in cell line supernatants.

Our results have important clinical implications. First, the theragnostic significance of *in vivo* Trp imaging, at least in MM, may justify the development of F^18^-based Trp PET probes for broader clinical use, given C11-AMT’s very short half-life. Such probes can be further designed to be “agnostic” or pathway-nonspecific (i.e., serotonin/melatonin vs. kynurenine), given that different cancers may preferentially metabolize Trp towards one pathway and not the other.^21^ Lastly, given the availability of TPH1 inhibitors in the clinic, targeting TPH1 in patients with PD1 inhibitor-refractory melanoma may potentially restore PD1 inhibitor resistance.

## Data Availability

All data produced in the present study are available upon reasonable request to the authors.

## Acknowledgements

We thank the patients and their families for participating in the LCCC1531 trial.

## Abbreviations

Trp: tryptophan
IDO1: indoleamine 2,3-dioxygenase
TDO2: tryptophan dioxygenase-2
MM: metastatic melanoma
TPH: tryptophan hydroxylase
TMEs: tryptophan-metabolizing enzymes
LAT1/SLC7A5: Large neutral amino acid transporter small subunit-1
TILs: tumor-infiltrating lymphocytes
OS: overall survival
CLA: cell line array
ICC: immunocytochemistry
TMA: tissue microarray
IHC: immunohistochemistry
SOC: standard of care
UNC-CH: University of North Carolina at Chapel Hill
PET: positron emission photography
FDG: fluorodeoxyglucose
CT: computerized tomography
IV: intravenous
RECIST, OR: odds ratio
95CI: 95% confidence intervals

## Funding Statement

This work was supported by The National Cancer Institute Cancer Clinical Investigator Team Leadership Award (5P30CA016086-38, SJM), NCI 1 R01 CA233904-01 (ZL and SJM), and Merck & Co., Inc (SJM and ZL).

## Disclosure of potential conflicts of interest

Funding for the LCCC1531 trial was provided by Merck & Co., Inc.

## Supplementary materials

Supplementary data for this article can be accessed on the publisher’s website.

## Data Availability

The data that support the findings of this study are available from the corresponding author, SJM, upon reasonable request.

## Notes

### Clinical Trial

NCT03089606

### Author Declarations

The study was approved by the UNC-CH IRB. Written informed consent was obtained from all patients in compliance with the IRB regulations.

