## Supplementary Material for "Increased Tryptophan, But Not Increased Glucose Metabolism, Predict Resistance of Pembrolizumab in Stage III/IV Melanoma"

### **Patients and Methods**

**Inclusion/Exclusion Criteria (LCCC1531 study)**

Eligible patients must have been at least 18 years old upon study enrollment, must have had Eastern Cooperative Oncology Group performance status 0-2, histologic or cytologic biopsy-proven diagnosis of stage IIIB-IV melanoma irrespective of histologic type (i.e., cutaneous, unknown primary, mucosal, or ocular), and measurable stage IIIB-IV melanoma using RECIST v1.1 criteria. In addition, patients should have had adequate organ function, including hematologic (hemoglobin ≥ 9 g/dL, neutrophil count ≥ 1,500 cells/μL, platelet count ≥ 100,000 cells/μL, prothrombin time ≤ 1.5x upper limits of normal [ULN], activated partial thromboplastin time ≤ 1.5x ULN) and hepatic (aspartate aminotransferase and alanine aminotransferase ≤ 5x ULN if liver metastases were present, or ≤ 2x ULN if absent, and ≤ 1.5x ULN unless Gilbert’s syndrome was suspected, albumin ≥ 2.5 mg/dL). Patients with resectable stage IIIB-D melanoma or prior treatment with ipilimumab or BRAF/MEK inhibitors were allowed as long as they had completely recovered from any toxicities that occurred during prior treatment.

Eligible patients should not have received prior treatment with PD-1/PD-L1 pathway inhibitors in the adjuvant setting. They should not have known active, symptomatic central nervous system metastases that required antiepileptic drugs or corticosteroids. Patients with previously treated brain metastases could participate provided they were stable (i.e., without evidence of progression by imaging) for at least two weeks prior to the first dose of pembrolizumab, and any neurologic symptoms must have returned to baseline. In addition, eligible patients should not have an active autoimmune disease that requires systemic treatments, non-infectious pneumonitis, or active infection that requires systemic therapy.

Eligible patients must have been willing to provide fresh research tumor tissue from an accessible tumor lesion prior to treatment and to be injected with α-[^11^C]-methyl-L-tryptophan (C11-AMT) prior to pembrolizumab treatment.

**Patient Monitoring during and after the study (LCCC1531 study)**

During the 12-week time interval of the study, we monitored toxicity by grading adverse events (AEs) using the Common Terminology Criteria for Adverse Events version 5.0. In addition, if patients had not progressed following four pembrolizumab infusions, they underwent SOC repeat whole body FDG-PET co-registered with CT scan of the N-C-A-P using IV contrast for response assessment.

Following study completion, patients were allowed to discuss the SOC treatment options with their physician. If patients chose to continue on pembrolizumab, we collected the following data: duration while on pembrolizumab, time-to-progression from pembrolizumab, type of treatments following pembrolizumab progression (if any), and status at last follow-up.

**C11-AMT PET imaging**

C11-AMT was manufactured at the Radiochemistry core (BG, ZL) of the Biomedical Research Imaging Center (BRIC), the University of North Carolina at Chapel Hill (UNC-CH), under an Investigation New Drug Application (IND129471). Radioisotope synthesis was carried out with 16.5 MeV protons produced using a General Electric PETTrace 880 cyclotron (GE Healthcare Systems, Chicago, IL). Carbon-11 carbon dioxide (^11^CO_2_) was generated via the ^14^N (p, α) ^11^C nuclear reaction using 99% N_2_/1% O_2_ gas (Airgas) as the target. ^11^CO_2_ was subsequently reduced and iodinated to form carbon-11 methyl iodide (^11^CH_3_I) using a GE Healthcare FX2C synthesis module.

AMT was synthesized using an adaptation of literature procedures.^1^ **Supplementary Figure 2** shows the two-step hydrolysis procedure we used to produce C11-AMT for this study. First, the AMT precursor (dimethyl 2(S), 3a(R), 8a(S)-(+)-hexahydro-8(phenylsulfonyl)pyrrolo[2, 3-b]indole-1,2-dicarboxylate, Aberjona Laboratories, Woburn, MA) was deprotonated by freshly prepared lithium diisopropylamide and alkylated with ^11^CH_3_I at -40°C. Then, the protecting groups were hydrolyzed with neat trifluoroacetic acid at 130°C and 5M potassium hydroxide at 160°C. The crude reaction material was neutralized with acetic acid and purified via semi-preparative high-pressure liquid chromatography using a 5% ethanol/95% phosphate buffer. The product was subjected to a series of quality tests to ensure that it was suitable for human administration, as shown in **Supplementary Table 1**. The Radiopharmacy and Quality Management for UNC’s Cyclotron and Radiochemistry Research Project released the product at the BRIC, UNC-CH (ES) solution as the mobile phase. The collected fraction was sterilized through a 0.22 µm filter.

**C11-AMT PET Scan Protocol**

We performed C11-AMT PET imaging using a Siemens Biograph mCT 64 PET/CT scanner (Siemens Medical Solutions, USA Inc, Malvern, PA). On the day of the scan, patients fasted for at least 6 hours. A low-dose scout CT scan (120 kVp, 10 mA) was initially acquired for patient positioning and determining dose modulation parameters. Based on this scout image, we selected a one-bed position and acquired a non-contrast, low-dose CT scan (120 kVp, 100 mA) for attenuation correction.

We administered a weight-based dose of C11-ΑΜΤ (0.1 mCi/kg) IV. 5 min following C11-AMT injection. We acquired a 40-min dynamic emission PET scan (20 x 120 secs) in high-sensitivity 3D mode. We generated PET data sets using the list-mode-based established approach in a one-bed position, and CT-based attenuation correction was applied. Each patient was scanned once, based on a previous report that showed no difference in AMT uptake when each subject was scanned twice.^2^

**Quantitative PET Image Analysis (C11-AMT PET and FDG PET)**

Following the acquisition, we reconstructed data using the Siemens-specific iterative TrueX+TOF reconstruction algorithm using three different images: first, dynamic images with/without attenuation correction (2 minutes per frame). Second, static images with/without attenuation correction for the time interval that corresponds to 5-10 minutes post-injection. Third, static attenuation-correction images for the time intervals that correspond to 10-20 minutes, 20-30 minutes, and 30-40 minutes post-injection.

We used the Ordered Subset Expectation Maximization algorithm for image reconstruction, with four iterations and 12 subsets. Images were 200 x 200 with a 5 mm slice thickness. For scatter correction, we used a model based on relative scatter scaling. Experienced observers (JO, YZL, TZW) visually identified tumors based on CT and FDG PET images. To objectively determine tumor regions of interest (ROI) on ^11^C-AMT and FDG PET, we initially determined the voxel with the highest AMT tracer concentration as well as a background region near the location of the tumor using the MIMvista PET viewing software (MIM Software Inc., Cleveland, OH, version 7.0.5). We then exported ROIs on static data to LIFEx (LIFEXsoft, Orsay, FR, version 5.10). Finally, we constructed a Logan plot for dynamic data on each lesion, excluding the first 10 min before equilibrium was reached.

#### In vitro studies

Melanoma cell lines (SK-MEL-2 and MeWo) were obtained from the American Type Culture Collection organization (Manassas, VA, USA). Cells were grown in Minimum Essential Medium (MEM) that contains 10% fetal bovine serum (FBS), 2 mM L-glutamine, 100 IU/ml penicillin, and 100 *μ*g/ml streptomycin (GIBCO, Gaithersburg, MD, USA), and cultured at 37°C in the incubator supplied with 5% CO_2_. Telotristat etiprate, the hippuric acid salt form of the telotristat ethyl, was generated after the prodrug, telotristat ethyl (#U105830, Advanced ChemBlocks Inc., Hayward, CA; dissolved in dimethylsulfoxide, DMSO), was mixed in equal amounts with hippuric acid (#112003, Sigma-Aldrich, Inc., St. Louis, MO; dissolved in water).

For Western blotting analysis, whole cell lysates were prepared from melanoma cell lines following treatment with telotristat etiprate for 24 hours. 20 mg of cell lysates were denatured and separated by SDS-PAGE. Separated proteins were then transferred onto a nitrocellulose membrane (Bio-Rad, Hercules, CA, USA). The membranes were blocked with 2% bovine serum albumin (BSA) in PBS for 1 hour and incubated overnight with antibodies against IDO1 (PA5-24598, Invitrogen, Waltham, MA; dilution 1:1,000), TPH1 (NBP1-86922, Novus Biologicals LLC, Centennial, CO; dilution 1:1,000), and β-actin (SC-69879, Santa Cruz Bioechnology Inc. Dallas, TX; dilution 1:5,000). Following incubation with corresponding IgG horseradish peroxidase-conjugated secondary antibodies (Cell Signaling Technology, Danvers, MA), immunoblots were developed using the enhanced chemiluminescence reagent (Cell Signaling) and visualized using an Imaging processor (Bio-Rad).

For the cell viability assay, 10,000–25,000 cells/well were plated in 24-well plates and incubated overnight. Cells were then treated with 1-10 mM telotristat etiprate for 72 hours. At the end of the follow-up period, MTT was added to the cultures and incubated for 4 hours at 37°C. Formed dark crystals were then dissolved by adding an equal volume of extraction buffer (20% SDS and 50% N, N-DMF [pH 4.7], in PBS). The absorbance of the soluble fraction was measured at 570 nm using an ELISA plate reader. Culture supernatants from melanoma cells treated with telotristat etiprate for 24 hours were used to measure kynurenine (LS-F39319, LSBio, Seattle, WA) and serotonin (ADI-900-175, Enzo Life Sciences, Farmingdale, NY).

#### Statistical Analysis

Melanocyte/Melanoma Cell line Array

We used descriptive statistics (mean and standard errors) to summarize protein expression levels (arbitrary fluorescence units above background, H-score) of TPH1, TPH2, TDO2, IDO1, and LAT1 across the NHM and melanoma cell lines. We performed an unpaired *t*-test to compare the expression among groups. We used ANOVA to compare more than two groups. If significant, we performed subsequent two-group comparisons using an unpaired *t-*test with Bonferroni corrections. We performed statistical analyses and developed graphs using Prism 8 (GraphPad Software, version 8.3.1, San Diego, CA, USA).

Normal Skin/Benign Nevi/Primary melanoma TMAs

We used descriptive statistics (mean and standard errors) to summarize protein expression levels (arbitrary fluorescence units above background, H-score) of TPH1, TPH2, TDO2, IDO1, and LAT1 in normal skin, nevi, and primary melanomas from the SK181, ME1002b commercially available TMAs. We used the Mann-Whitney test to compare the expression of each biomarker between nevi and primary melanomas. We performed statistical analyses and developed graphs using Prism 8 (GraphPad).

#### Melanoma Tissue Microarray (09-1737 cohort)

We converted each expression category (0, 1, 2, 3) to the midpoint of the expression interval (i.e., category 0 corresponds to 0, category 1 to 12.5, category 2 to 52.5, and category 3 to 90). Within each sample and each compartment (i.e., melanoma, TILs), we averaged expression based on the number of available replicates. We used paired *t*-tests to compare the average expression of each protein in each melanoma and TIL compartments; and second, TPH1, TPH2, TDO2, and LAT1 in melanoma cells with that of IDO1 in melanoma cells.

To assess the relationship between the expression of tryptophan-metabolizing enzymes and LAT1 with the status of TILs, we fit a generalized linear mixed model with logit link function for each protein allowing for a random intercept by specimen case. This model estimates the odds of tumor invasion by lymphocytes for each expression level of the five proteins, adjusting for the correlation within replicates from the same specimen. The model fitted provides the estimated effect of a unit increase of tumor expression (on the 0-100 scale) on the log odds of tumor invasion by lymphocytes. The exponentiated coefficients correspond to the multiplicative effect of each unit increase in expression on the odds of lymphocyte invasion. We used the Wald test to determine if expression levels of each protein were significantly associated with TIL presence.

To evaluate whether expression levels of any of the five-tryptophan pathway-associated proteins predict melanoma-specific OS, we fit a Cox proportional hazards model for each of the five proteins to model OS based on average tumor expression. This model included staging information (III or IV), age, and sex as covariates. We treated patients who died from causes other than melanoma as censored. We used the Wald test to investigate the association of each covariate with OS.

#### Clinical Trial (LCCC1531)

We used descriptive statistics to present demographics, melanoma subtypes, stage at study enrollment, SUV_max_ of each tumor imaged by both C11-AMT PET and FDG PET scan at baseline, duration of pembrolizumab treatment, the best antitumor response by RECIST v.1.1, treatment-emergent adverse events, progression-free survival (PFS) while on pembrolizumab, and melanoma-specific overall survival (OS). For the PFS endpoint, in particular, we censored patients who elected to have definitive surgery after the four pembrolizumab infusions administered as part of the LCCC1531 study. In addition, we performed survival analysis to assess the prognostic significance of the patient’s tumor exhibiting the highest maximum standardized uptake value (SUV_max_) for the C11-AMT PET and FDG PET scan at baseline (i.e., before pembrolizumab treatment) from the LCCC1531 study. We fitted a Cox proportional hazards model for PFS and OS with baseline SUV_max_ C11-AMT PET or baseline SUV_max_ FDG PET imaging as a continuous variable. As a second approach, we performed exploratory analysis using the Kaplan-Meier method. In particular, we used the optimal cut-point method for a ‘high’ versus ‘low’ SUV_max_ for the C11-AMT PET scan and FDG PET scan at baseline, as we have previously described.^3,4^ The clinical question was whether patients who remained free of recurrence while on pembrolizumab treatment or lived longer demonstrated ‘higher’ or ‘lower’ SUV_max_ for C11-AMT and FDG-PET prior to pembrolizumab and for a given cut-point. We performed statistical analyses and developed graphs using Prism 8 (GraphPad). Results were considered statistically significant if the two-tailed *p*-value was ≤ 0.05. To assess whether a given cut-point baseline SUV_max_ for C11-AMT (or FDG-PET) was associated with progression vs. non-progression (the sum of complete response, partial response, and stable disease) while on pembrolizumab by RECIST v1.1 criteria, we used a two-tailed Fisher’s exact test.

We used descriptive statistics to compare the expression of the eight proteins by melanoma cells and by TILs if present. To assess whether TIL density or the melanoma-specific expression of any of the eight tryptophan and glucose metabolism-associated proteins at baseline (i.e., prior to pembrolizumab) was associated with PFS or OS benefit, we dichotomized the density (or expression) as absent/non-brisk (0, 1+) versus brisk (2+, 3+), as we have previously published.^3-5^ To assess whether, for a given cut-point, baseline SUV_max_ for C11-AMT (or FDG-PET) was associated with absent/non-brisk versus brisk expression of any of the tryptophan- or glucose-pathway associated proteins, respectively, we used a two-tailed Fisher’s exact test.

We used the Kendall tau rank correlation statistic to explore the association between baseline C11-AMT and FDG PET SUV_max_ values of biopsied tumors and melanoma cell-specific expression of IDO1 in these PET-imaged tumors. In addition, we used the Spearman rank correlation statistic to explore the association in protein expression among various tryptophan and glucose-metabolizing enzymes in melanoma cells and TILs.

Supplementary Figures

**Supplementary Figure 1. Treatment schema for the LCCC1531 prospective clinical trial (**clinicaltrials.gov, NCT03089606**), “***Pembrolizumab in Systemic Treatment-Naïve Metastatic Melanoma and Exploration of Use of Baseline ^11^C-methyl-L-tryptophan (C11-AMT) PET Imaging as a Predictive Imaging Biomarker of Antitumor Response*.” *Abbreviations*: FDG, fluoro-deoxyglucose; PET, positron emission tomography; CTIV, computerized tomography scan with intravenous contrast; pembro, pembrolizumab; IV, intravenously.

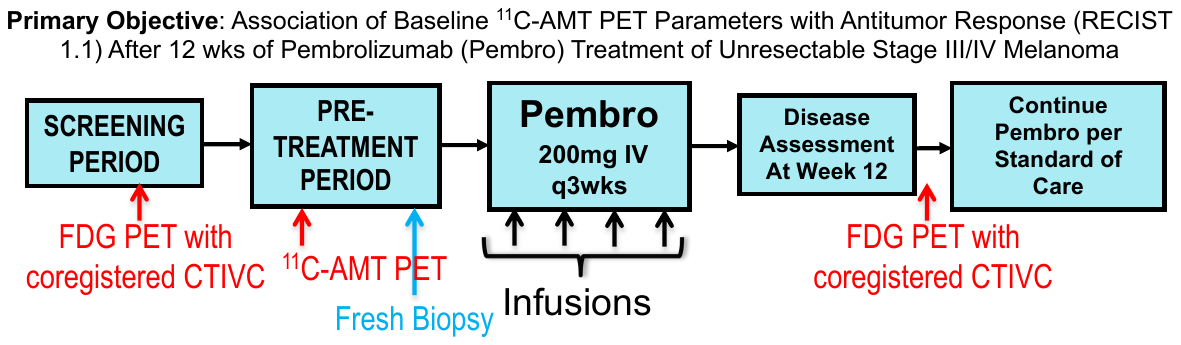

**Supplementary Figure 2. Procedure for synthesis of Carbon-11 α-Methyl-L-Tryptophan.** Abbreviations: LDA: lithium diisopropylamide; THF, tetrahydrofuran; TFA, trifluoroacetic acid; C-18 Sep-Pak, solid phase extraction chromatography cartridges. Adapted from.^6^

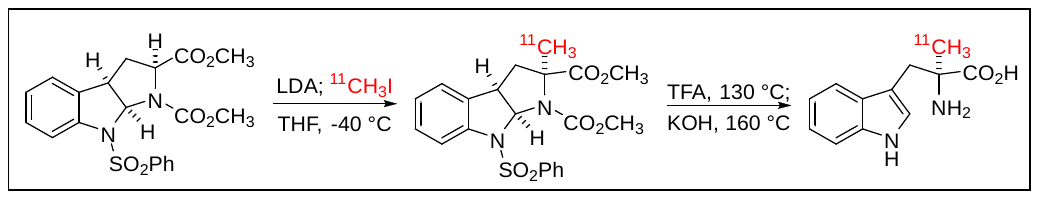

**Supplementary Figure 3. Protein expression of tryptophan pathway (Trp, light blue and dark blue) and glucose pathway (Glc, light and dark green) components in PD1 inhibitor-naïve metastatic melanoma.** Expression is measured using the 0, 1+, 2+, 3+ scale and for melanoma cells (mel) and tumor-infiltrating lymphocytes (TILs, if present) separately. See Materials and methods for details. Numbers above each violin plot indicate the number of patients. Asterisks indicate Bonferroni-adjusted significant differences in expression between the two tissue compartments.

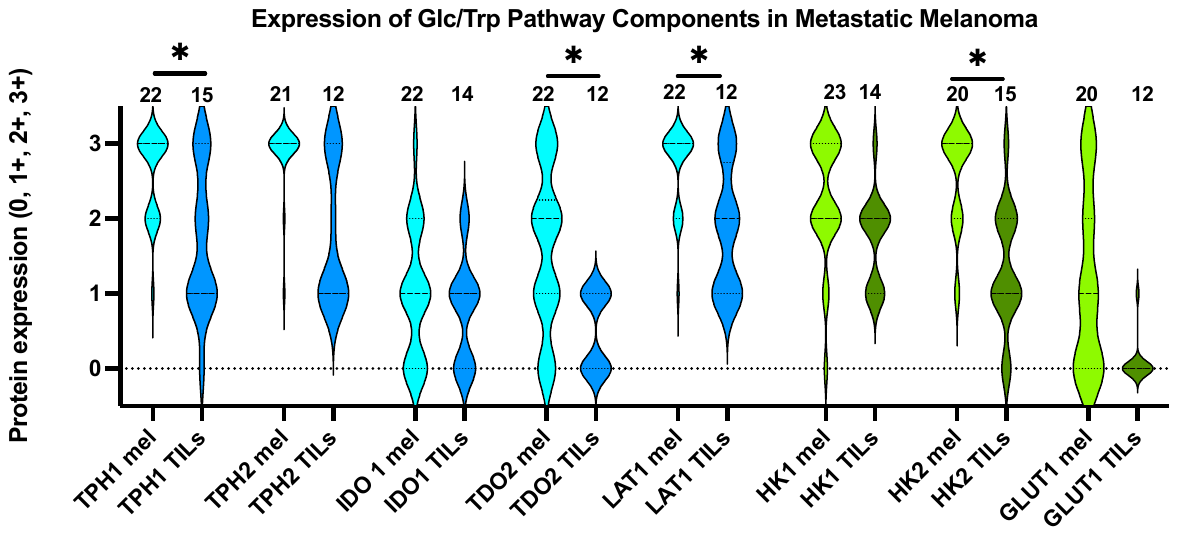

Supplementary Tables

**Supplementary Table 1.** **Summary of quality control tests required prior to the release of the radiopharmaceutical product for human studies.** Abbreviations: USP, United States Pharmacopeia; HPLC, high-pressure liquid chromatography; TLC, thin layer chromatography; Rf, relative front (a measure of solubility); ppm, parts per million; EU, endotoxin units.

| **Quality control test** | **Description** | **Requirements for pass** | **Requires test pass prior to product release** |
| --- | --- | --- | --- |
| **Chemical Purity** | Visual inspection for  color and particulates | Clear and Colorless | yes |
| **Filter Integrity** | Test bubble point | Meet pressure specified by manufacturer | yes |
| **pH** | pH as per USP test method 791 | 5.0 – 7.5 | yes |
| **Chemical & Radiochemical Purity** | HPLC consistent with guidelines of General Chapter <621>, Chromatography subsection HPLC | Radiochemical Purity > 95%  Other UV (list nm) peaks ≤ 35 µg in final product | yes |
| **Radiochemical Purity** | TLC | Rf > 0.5 and  Purity ≥ 95% | yes |
| **Residual Solvent Levels** | Gas Chromatography | Acetone < 5,000 ppm  Acetonitrile < 400 ppm | yes |
| **Radionuclidic Purity** | Half-life Determination | 18.4 – 22.4 min | yes |
| **Bacterial Endotoxin Levels** | Limulus Amoebocyte Lysate (LAL) by PTS | < 175 EU per dose | yes |
| **Sterility** | USP <71> sterility test | No growth observed in 14 days | no |

**Supplementary Table 2.** **Antibody characteristics and details regarding single-color immunohistochemistry staining protocol.** For each of the eight antibodies used for this study. *Abbreviations*: HIER, heat-induced epitope retrieval; *, incubation occurs at room temperature (RT); P, polyclonal; M, monoclonal; Cat #, catalog number; Dil, dilution; hrs, hours; temp, incubation temperature; O/N, overnight; RT, room temperature; min, minutes; AP, alkaline phosphatase; ABC, three-step detection immunohistochemistry method.

| **Protein** | **Antigen Retrieval** | | **Protein Block** (min) | **Primary Antibody** | | | | **Secondary Antibody** | | | **Signal Detection (min)** | |
| --- | --- | --- | --- | --- | --- | --- | --- | --- | --- | --- | --- | --- |
|  | Buffer type (min) | Vendor  (Cat #) |  | Clonality | Vendor (Cat #) | Dil (1:x) | Time/Temp (hrs-ºC) | Kit (Cat #) | Time (min) | Vendor | Kit (min) | Vendor |
| TPH1 | HIER L* | Thermo Fisher Scientific  (TA-135-HBL) | 10% normal goat serum (60)* | Rabbit P | Novus Biologicals (NBP1-86922) | 150 | O/N-4 | Biotinylated goat anti-rabbit | 60* | Jackson ImmunoResearch (111-065-144) | ABC-AP (30)* with Immpact Vector Red (Vector Lab SK-5105) | Vector Lab (AK5000) |
| TPH2 |  |  |  | Rabbit P | Novus Biologicals  (NB100-74555) | 150 |  |  |  |  |  |  |
| IDO-1 |  |  |  | Mouse M | Thermo Fisher Scientific (TF14-9750-82) | 200 |  | Biotinylated goat anti-mouse |  | Jackson ImmunoResearch (111-605-166) |  |  |
| LAT-1 |  |  |  | Rabbit P | Abcam (Ab85226) | 200 |  | Biotinylated goat anti-rabbit |  | Jackson ImmunoResearch (111-065-144) |  |  |
| TDO2 | Borg Decloaker (10)* | Biocare  (BD1000) | DAKO Z0909 (10)* | Mouse M | Millipore Sigma (MABN1537) | 50 |  | MACH4 Universal AP Probe | 15* | Biocare (UP536H) | MACH4 MR-AP Polymer (20)* with Immpact Vector Red (Vector Lab SK-5105) | Biocare (MRAP536H) |
| Glut1 | HIER 1 (20)* | Leica Biosystems  (AR9961) | No | Mouse M | Biocare (CM408A) | 200 | 60-RT | BOND Polymer Refine Red Detection | 30* | Leica Biosystems (D59390) | Bond Polymer Refine Red Detection AW-Red (60)* | Leica Biosystems (D59390) |
| HK1 |  |  |  | Rabbit M | Cell Signaling (2024S) | 800 |  |  |  |  |  |  |
| HK2 |  |  | Background Sniper (Biocare, BS966MM) (10)* | Mouse M | Abcam (ab104836) | 300 | 30-RT |  |  |  | Bond Polymer Refine Red Detection AW-Red (30)* |  |

**Supplementary Table 3. Expression of the four tryptophan-metabolizing enzymes and the tryptophan transporter, LAT1, in normal human melanocytes (NHM) and various melanoma cell lines.** Each cell line was spotted in the cell line array in triplicates. Results are shown as H-score means with standard deviations. ‘*No aberrations’* is defined as no genetic aberrations for *BRAF*, *NRAS*, *TP53*, *MSH6*, *EPH6*, *APC*, *CDKN2A*, *ATM*, *PTEN*, *MTOR*, *MET*, *FLT1*, *ERBB4*, *STK11*, *RB1*, *PIK3CA*, *KRAS*, *KIT*, *HRAS* by exome sequencing.

| **Cell Lines** |  | **TPH1** | **TPH2** | **TDO2** | **IDO1** | **LAT1** |
| --- | --- | --- | --- | --- | --- | --- |
| NHM 7 |  | 121 ± 3 | 26 ± 19 | 1.1 ± 1.4 | 83 ± 22 | 107 ± 15 |
| NHM 11 |  | 149 ± 5 | 91 ± 16 | 0.4 ± 0.3 | 129 ± 23 | 146 ± 7 |
| NHM 12 |  | 153 ± 5 | 101 ± 10 | 0.7 ± 0.4 | 131 ± 28 | 156 ± 7 |
| 1205Lu |  | 161 ± 11 | TDO | 1.9 ± 2.6 | 154 ± 6 | 158 ± 7 |
| A2058 | *BRAF*^V600^, *TP53*^mut^, *APC*^mut^, *ATM*^mut^, *MTOR*^mut^, *FLT1*^mut^, *ERBB4*^mut^, *STK11*^mut^, *PTEN*^mut^ | 150 ± 8 | 44 ± 19 | 2.1 ± 1.4 | 125 ±18 | 149 ± 13 |
| A375 | *BRAF*^V600^, *CDKN2A*^loss^ | 145 ± 15 | 58 ± 19 | 1.4 ± 0.5 | 116 ± 6 | 142 ± 7 |
| Malme 3 | *BRAF*^V600,^ *CDKN2A*^loss^ | 150 ± 8 | 75 ± 19 | 8.4 ± 13 | 131 ± 15 | 154 ± 3 |
| mel 224 | *NRAS*^Q61^, *MSH6*^mut^ | 153 ± 4 | 48 ± 9 | 0.5 ± 0.3 | 120 ± 17 | 145 ± 5 |
| mel 505 | No aberrations | 150 ± 4 | 72 ± 6 | 2.9 ± 1.9 | 117 ± 22 | 152 ± 3 |
| mel 537 | *BRAF*^V600^, *PTEN*^loss^, *CDKN2A*^loss^, *EPHA6*^mut^, *ATM*^mut^, *MTOR*^mut^, *FLT1*^mut^ | 140 ± 22 | 58 ± 8 | 4.8 ± 4.5 | 96 ± 27 | 143 ± 15 |
| PMWK | *PTEN*^loss^ | 158 ± 6 | 90 ± 12 | 2.7 ± 1.5 | 113 ± 30 | 158 ± 13 |
| RPMI7951 | *BRAF*^V600^, *PTEN*^loss^, *TP53*^mut^, *MET*^mut^ | 161 ± 12 | 76 ± 5 | 0.8 ± 0.4 | 152 ± 7 | 162 ± 1 |
| RPMI8322 | *KRAS*^mut^, *APC*^mut^, *TP53*^mut^ | 138 ± 39 | 96 ± 14 | 4.9 ± 6.3 | 124 ± 19 | 134 ± 26 |
| SBC12 | *NRAS*^Q61^, *APC*^mut^, *ATM*^mut^ | 150 ± 1 | 30 ± 3 | 1.5 ± 1.2 | 128 ± 18 | 150 ± 6 |
| SKMEL5 | No aberrations | 155 ± 8 | 50 ± 6 | 0.8 ± 0.4 | 145 ± 8 | 148 ± 5 |
| SKMEL23 | *BRAF*/*NRAS*^wt^, *BRAF*^fusion^, *PTEN*^loss^, others unkn | 157 ± 1 | 119 ± 13 | 4.8 ± 7.9 | 139 ± 8 | 154 ± 1 |
| SKMEL24 | *BRAF*^V600^, *PTEN*^loss^, *CDKN2A*^loss^ | 146 ± 13 | 86 ± 5 | 0.9 ± 0.3 | 137 ± 7 | 146 ± 7 |
| SKMEL27 | *BRAF*^V600^, others unkn | 155 ± 6 | 82 ± 8 | 1.0 ± 0.5 | 124 ± 17 | 146 ± 8 |
| SKMEL28 | *BRAF*^V600^, *PTEN*^mut^ | 146 ± 2 | 50 ± 3 | 0.3 ± 0.3 | 125 ± 10 | 140 ± 2 |
| SKMEL78 | *HRAS*^mut^, *TP53*^mut^ | 133 ± 4 | 44 ± 8 | 2.0 ± 0.6 | 97 ± 20 | 134 ± 9 |
| SKMEL88 | *HRAS*^mut^, *TP53*^mut^ | 125 ± 12 | 36 ± 12 | 1.3 ± 0.5 | 86 ± 15 | 124 ± 11 |
| SKMEL100 | *BRAF*^V600^, *TP53*^mut^, *CDKN2A*^loss/mut^, *APC*^mut^, *ATM*^mut^ | 146 ± 21 | 67 ± 10 | 11 ± 6 | 110 ± 19 | 148 ± 23 |
| SKMEL103 | *NRAS*^Q61^, others unkn | 154 ± 14 | 80 ± 15 | 4.5 ± 1.7 | 133 ± 3 | 150 ± 6 |
| SKMEL115 | *BRAF*^V600^, others unkn | 161 ± 18 | 64 ± 26 | 11 ± 6 | 144 ± 8 | 170 ± 11 |
| SKMEL119 | *NRAS*^Q61^, *CDKN2A^l^*^oss^, *EPHA6*^mut^ | 136 ± 18 | 41 ± 16 | 2.2 ± 3.2 | 115 ± 7 | 134 ± 11 |
| SKMEL130 | *BRAF*^V600^ | 160 ± 14 | 72 ± 4 | 4.2 ± 3.8 | 132 ± 7 | 152 ± 13 |
| SKMEL131 | No aberrations | 143 ± 6 | 27 ± 18 | 1.1 ± 1.5 | 141 ± 11 | 137 ± 12 |
| SKMEL147 | *NRAS*^Q61^, *CDKN2A*^loss/mut^, *MSH6*^mut^, *ERBB4*^mut^ | 142 ± 6 | 72 ± 18 | 1.3 ± 1.1 | 110 ± 13 | 135 ± 7 |
| SKMEL153 | *BRAF*^V600^, *EPHA6*^mut^ | 170 ± 3 | 68 ± 10 | 26 ± 18 | 119 ± 21 | 168 ± 4 |
| SKMEL173 | *NRAS*^Q61^, *CDKN2A*^loss^, *EPHA6*^mut^, *MET*^mut^ | 148 ± 3 | 61 ± 7 | 1.3 ± 0.7 | 106 ± 20 | 150 ± 5 |
| SKMEL178 | *BRAF*^V600^, *PTEN*^loss^, others unkn | 154 ± 5 | 66 ± 13 | 1.4 ± 0.9 | 134 ± 12 | 148 ± 5 |
| SKMEL181 | *BRAF*^V600^, *CDKN2A*^loss^, *EPHA6*^mut^, *APC*^loss^ | 162 ± 8 | 117 ± 7 | 1.2 ± 0.5 | 122 ± 11 | 164 ± 4 |
| SKMEL186 | *CDKN2A*^mut^, *TP53*^mut^, *MSH6*^mut^ | 148 ± 14 | 53 ± 27 | 0.3 ± 0.2 | 119 ± 26 | 150 ± 15 |
| SKMEL187 | *CDKN2A*^loss^, *TP53*^mut^, *MSH6*^mut^ | 153 ± 41 | 101 ± 48 | 2.9 ± 1.3 | 143 ± 33 | 151 ± 37 |
| SKMEL190 | *BRAF*^V600^, *PTEN*^loss^, *CDKN2A*^loss^, *ATM*^mut^ | 141 ± 2 | 47 ± 29 | 1.1 ± 1.2 | 135 ± 15 | 145 ± 9 |
| SKMEL235 | *BRAF*^V600^, *PTEN*^loss^, *CDKN2A*^loss^ | 146 ± 3 | 29 ± 16 | 0.4 ± 0.1 | 129 ± 15 | 145 ± 5 |
| SKMEL239 | *BRAF*^V600^, others unkn | 152 ± 6 | 63 ± 23 | 0.4 ± 0.1 | 135 ± 11 | 151 ± 6 |
| UACC 257 | *BRAF*^V600^, *MET*^mut^ | 170 ± 11 | 78 ± 24 | 8.3 ± 4.4 | 130 ± 9 | 170 ± 3 |
| VMM39 | *NRAS*^Q61^ | 153 ± 8 | 42 ± 17 | 2.4 ± 0.9 | 125 ± 9 | 151 ± 8 |
| WM35 | *BRAF*^V600^, others unkn | 158 ± 9 | 101 ± 14 | 2.0 ± 1.5 | 142 ± 8 | 162 ± 8 |
| WM1158 | *BRAF*^V600^, *PTEN*^mut^, *ATM*^mut^, *MET*^mut^ | 140 ± 21 | 58 ± 20 | 0.1 ± 0.1 | 109 ± 44 | 145 ± 6 |
| WM1232 | *BRAF*^V600^, *TP53*^mut^, *MSH6*^mut^, *CDKN2A*^mut^, *ATM*^mut^ | 166 ± 12 | 115 ± 32 | 1.8 ± 1.3 | 148 ± 9 | 166 ± 6 |
| WM2664 | *BRAF*^V600^, *PTEN*^loss^, *CDKN2A*^loss^, *MET*^mut^ | 152 ± 8 | 59 ± 22 | 3.2 ± 2.6 | 130 ± 17 | 163 ± 7 |
| WM1361A | *NRAS*^Q61^ | 133 ± 38 | 79 ± 10 | 0.2 ± 0.0 | 144 ± 2 | 156 ± 6 |

**Supplementary Table 4.** Patient characteristics of the 09-1737 tumor tissue cohort (n=87)

| **Age at Original Diagnosis (years)** | |
| --- | --- |
| 21–40 | 20 (23) |
| 41–60 | 26 (30) |
| 61–80 | 37 (42) |
| >80 | 4 (5) |
| **Sex** | |
| Male | 59 (68) |
| Female | 28 (32) |
| **Ethnicity** | |
| White | 84 (97) |
| Non-Hispanic or Latino | 82 (95) |
| Hispanic or Latino | 2 (2) |
| African American | 2 (2) |
| Asian | 1 (1) |
| **Histology** | |
| Superficial spreading | 23 (26) |
| Nodular | 13 (15) |
| Non-superficial spreading, non-nodular | 23 (26) |
| Not otherwise specified | 29 (33) |
| **Site Origin** | |
| Head and neck | 29 (33) |
| Trunk | 27 (30) |
| Extremity | 26 (30) |
| Not otherwise specified | 6 (7) |
| **Breslow thickness** | |
| <1 mm | 10 (11) |
| 1–4 mm | 43 (49) |
| >4 mm | 24 (27) |
| Not otherwise specified | 11 (13) |
| **Ulceration** | |
| Present | 30 (34) |
| Absent | 28 (32) |
| Not otherwise specified | 30 (34) |
| **TIL Presentation** | |
| Brisk | 2 (2) |
| Non-Brisk | 37 (42) |
| Absent | 14 (16) |
| Not otherwise specified | 35 (40) |
| **Mitotic Rate** | |
| <1 | 15 (17) |
| ≧1 | 44 (50) |
| Not otherwise specified | 29 (33) |

**Supplementary Table 5.** Estimated Odds Ratio of TIL status in stage III/IV melanoma tumors for a unit increase in the average protein expression of the five principal components of the tryptophan metabolizing pathway using a generalized estimating equation. See text for details.

|  | **Log OR Estimate** | **OR Estimate** | **Std. Error** | **Z-value** | **P-value** | **95% CI** |
| --- | --- | --- | --- | --- | --- | --- |
| **TPH1** | -0.018 | 0.982 | 0.008 | -2.174 | 0.030 | 0.967,0.998 |
| **TPH2** | -0.003 | 0.997 | 0.007 | -0.383 | 0.702 | 0.984,1.011 |
| **IDO1** | 0.014 | 1.014 | 0.009 | 1.478 | 0.139 | 0.996,1.033 |
| **TDO2** | -0.009 | 0.991 | 0.006 | -1.374 | 0.169 | 0.979,1.004 |
| **LAT1** | -0.016 | 0.984 | 0.007 | -2.279 | 0.023 | 0.970,0.998 |

**Supplementary Table 6.** Estimated hazard ratio (HR) using a Cox proportional hazards model that includes average *melanoma cell-specific expression* of all 5 components of the tryptophan pathway, stage (III vs. IV), age, and sex (n=87).

| **Melanoma cell-specific protein expression** | **logHR Estimate** | **HR Estimate** | **Std. Error (logHR)** | **z** | **Pr(>⏐z⏐)** |
| --- | --- | --- | --- | --- | --- |
| TPH1 | 0.013 | 1.014 | 0.009 | 1.499 | 0.134 |
| TPH2 | -0.014 | 0.986 | 0.010 | -1.355 | 0.175 |
| **IDO1** | -0.021 | 0.979 | 0.010 | -2.204 | **0.028** |
| TDO2 | -0.005 | 0.995 | 0.006 | -0.853 | 0.393 |
| **LAT1** | 0.018 | 1.018 | 0.007 | 2.416 | **0.016** |
| **Stage IV** | 1.204 | 3.333 | 0.296 | 4.064 | **<0.0001** |
| Age | -0.010 | 0.990 | 0.008 | -1.229 | 0.219 |
| Male | -0.477 | 0.621 | 0.298 | -1.599 | 0.110 |

**Supplementary Table 7. Adverse events in the LCCC1531 trial.** Seventy-five adverse events (AEs) unlikely, possibly, probably, or definitely attributed to either C11-AMT or pembrolizumab were reported in 23 study patients during the 12-week study period. Of these, only four (all grade 1) were attributed to the C11-AMT infusion (dysesthesia, dysgeusia, AST/ALT elevation). Only two AEs were grade 3 (a grade 3 retinal detachment and a grade 3 reduction in neutrophil count). Summary of the most frequent (i.e., more than one study subject) treatment-related adverse events.

| **Adverse Event** | **Number (%) Study subjects** |
| --- | --- |
| Fatigue | 21 (81) |
| Pruritus | 5 (19) |
| Maculopapular rash | 4 (15) |
| Arthralgia | 4 (15) |
| Hyperthyroidism | 4 (15) |
| Serum ALT elevation | 3 (12) |
| Serum AST elevation | 3 (12) |
| Dry mouth | 3 (12) |
| Arthritis | 2 (8) |
| Serum total bilirubin elevation | 2 (8) |
| Blurred vision | 2 (8) |
| Serum creatinine increase | 2 (8) |
| Diarrhea | 2 (8) |
| Headache | 2 (8) |
| Hot flashes | 2 (8) |
| Hypothyroidism | 2 (8) |
